# Genomic and multi-tissue proteomic integration for understanding the biology of disease and other complex traits

**DOI:** 10.1101/2020.06.25.20140277

**Authors:** Chengran Yang, Fabiana G. Farias, Laura Ibanez, Brooke Sadler, Maria Victoria Fernandez, Fengxian Wang, Joseph L. Bradley, Brett Eiffert, Jorge A. Bahena, John P. Budde, Zeran Li, Umber Dube, Yun Ju Sung, Kathie A. Mihindukulasuriya, John C. Morris, Anne Fagan, Richard J. Perrin, Bruno Benitez, Herve Rhinn, Oscar Harari, Carlos Cruchaga

**Author notes:** To whom correspondence should be addressed: Carlos Cruchaga, PhD, Washington University, School of Medicine, 425 S. Euclid Ave., BJC Institute of Heath. Box 8134, St. Louis, MO 63110. Authors with the same contribution (Oscar Harari, Carlos Cruchaga).

## Abstract

Expression quantitative trait loci (eQTL) mapping has successfully resolved some genome-wide association study (GWAS) loci for complex traits^1–6^. However, there is a need for implementing additional “omic” approaches to untangle additional loci and provide a biological context for GWAS signals. We generated a detailed landscape of the genomic architecture of protein levels in multiple neurologically relevant tissues (brain, cerebrospinal fluid (CSF) and plasma), by profiling thousands of proteins in a large and well-characterized cohort. We identified 274, 127 and 32 protein quantitative loci (pQTL) for CSF, plasma and brain respectively. We demonstrated that cis-pQTL are more likely to be shared across tissues but trans-pQTL are tissue-specific. Between 78% to 87% of pQTL are not eQTL, indicating that protein levels have a different genetic architecture than gene expression. By combining our pQTL with Mendelian Randomization approaches we identified potential novel biomarkers and drug targets for neurodegenerative diseases including Alzheimer disease and frontotemporal dementia. In the context of personalized medicine, these results highlight the need for implementing additional functional genomic approaches beyond gene expression in order to understand the biology of complex traits, and to identify novel biomarkers and potential drug targets for those traits.

---

Genetic studies have been successful in identifying genetic regions associated with complex traits, including diabetes, cardiovascular disease and neurodegenerative diseases among others^1–4^, but have fallen short in understanding the biological mechanisms underlying those traits. Most genome-wide association studies (GWAS) identify multiple disease loci rather than the functional variant or genes, which makes it difficult to biologically interpret the association results and identify novel biomarkers and drug targets. By leveraging gene-expression and genetic data generated by multiple studies^5,6^, including the Genotype-Tissue Expression (GTEx) project, it has been possible to identify the functional variants or genes driving some GWAS signals. GTEx and others have shown that there are tissue-specific expression quantitative trait loci (eQTL), and that in order to perform appropriate eQTL mapping, it is important to interrogate the tissue of interest for the specific trait in question.

However, eQTL mapping has not been able to resolve all GWAS signals. There is evidence that many genetic variants alter protein and not transcript levels^7^. There are several published studies analyzing the genetic architecture of protein levels. Most are focused on a single tissue, mainly plasma^7–10^, though there are few studies with small sample sizes using cerebrospinal fluid^11,12^ and brain tissue^13^. These studies suggest that a good proportion of the protein QTL (pQTL) are not eQTL, and that additional GWAS signals can be resolved with pQTL mapping. Integration of pQTL with Mendelian Randomization has led to the identification of causal pathway and novel biomarkers for complex traits as well as compounds that could be used for drug repurposing^7^.

In this study, we combined high-throughput proteomics in multiple tissues with genetic data to determine the genomic architecture of protein levels in neurologically relevant tissues (brain, CSF and plasma), leading to the identification of multi-tissue and tissue-specific pQTL that are critical for the understanding of the biology of complex traits and diseases, mainly for CNS-related traits including neurodegenerative and psychiatric diseases.

## Multi-tissue pQTL

We generated data for 1,305 proteins using an aptamer-based approach in CSF (n=971), plasma (n=636) and brain (n=459) samples (**Extended Fig**.**1, Table S1**). We included multiple technical and biological replicates to confirm the replicability and reproducibility of our proteomic measurements (**Extended Fig**.**2**). We performed stringent quality control (QC) steps for the proteomic data (See Material and Methods). After QC, 835 CSF samples and 713 proteins, 529 plasma samples and 931 proteins, and 380 brain samples and 1079 proteins were included in the analyses (**Table 1 and S2**). To identify pQTL within each tissue (**Fig. 1a**), we performed genome-wide association analyses of 14.06 million imputed autosomal common variants (minor allele frequency (MAF) ≥ 0.02) against protein levels in each tissue. We defined cis-signals as when the single nucleotide polymorphism (SNP) fell within 1Mb upstream or downstream of the coding region, and trans-signals as when the single SNP fell outside of that window. To correct for multiple-tests, we used a stringent genome-wide threshold of p<5×10^−8^ for cis-pQTL and 5×10^−8^/number of independent proteins for trans-pQTL. There were 169, 230 and 75 independent proteins that passed QC in CSF, plasma and brain respectively (see Material and Methods), therefore the p-value thresholds were set at 2.96×10^−10^ for CSF; 2.17×10^−10^ for plasma, and 6.67×10^−10^ for brain.

**Table 1.** Demographics of the baseline cohort.

|  | **CSF** | **Plasma** | **Brain** |
| --- | --- | --- | --- |
| **N** | 835 | 529 | 380 |
| Age [mean+/-sd] | 69.4+/-9.3 | 69.8+/-9.4 | 83.3+/-10 |
| Female (%) | 53% | 54% | 57% |
| % CDR=0 | 74.37% | 68.24% | 11.57% |
| APOE e4 (%) | 38% | 41% | 48% |

**Fig 1.**
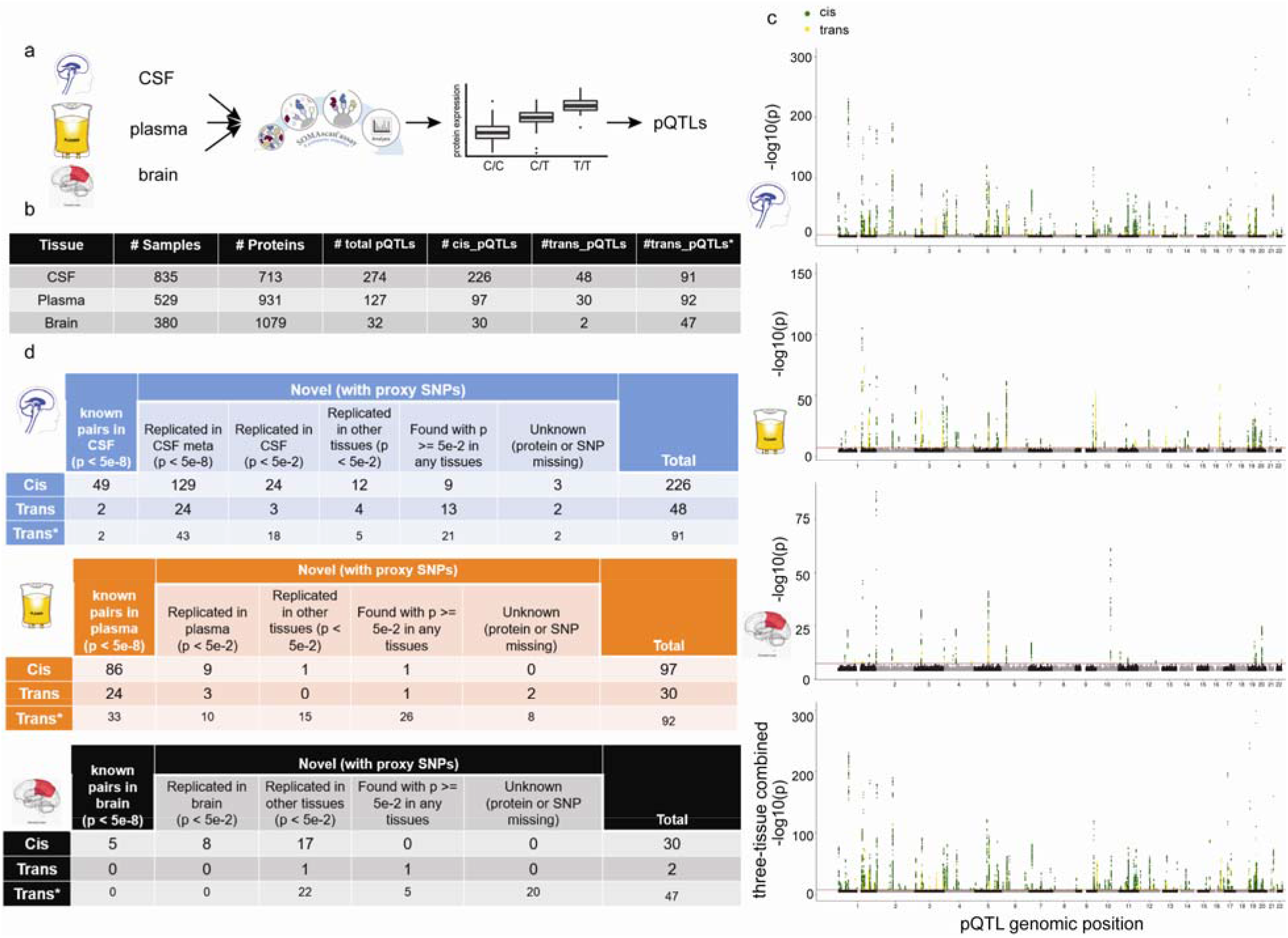
Study design and overview of the genome-wide associations of protein levels within each tissue. **(a**) Schematic of study design. CSF, plasma, brain tissues were profiled using a high-throughput aptamer-based proteomics platform. We identified common genetic variation within each tissue associated with each protein after integrating both the genotype for each variant and protein level. (**b**) Table of sample size after QC and total number of pQTL (split by cis, P < 5×10^−8^, and trans P < 5×10^−8^/number_PCs) for each tissue. For trans-pQTL, the p-value cutoff for CSF is 3×10^−10^ (5×10^−8^/169), for plasma it is 2×10^−10^ (5×10^−8^/230) and for brain it is 7×10^−10^ (5×10^−8^/75). Trans* represents replication of trans-pQTL given genome-wide significance (P-value < 5×10^−8^). **(c)** Stacked-Manhattan-plots for all three tissues mapping genomic locations of these pQTL within each tissue (cis: dark-green; trans: gold). The X-axis denotes the positions of the common variants. (**d**) Tables of replication of these pQTL within CSF, plasma and brain, given different nominal P-value thresholds on different datasets. Overall, we classified pQTL into five mutually exclusive groups: 1) known pQTL in the matched-tissue (single-study) with a p-value less than 5×10^−8^; 2) replicated pQTL in the matched-tissue with a p-value less than 5×10^−2^ but greater than or equal to 5×10^−8^; [*NOTE: for CSF, we split this group into two sub-groups: 2a) replicated only in the meta-analysis of two external CSF studies with a p-value less than 5×10^−8^; 2b) replicated pQTL in the matched-tissue with a p-value less than 5×10^−2^ but greater than or equal to 5×10^−8^] 3) replicated ones in the other tissues with a p-value less than 5×10^−2^; 4) found in any tissues (matched or not) with a p-value greater than or equal to 5×10^−2^; 5) unknown (either protein or SNP missing). For CSF, we further split the 2nd group into 2a) replicated pQTL in the matched-tissue (meta-analysis, Table S6) with a p-value less than 5×10^−8^ and 2b) replicated pQTL in the matched-tissue (meta-analysis and/or single-study) with a p-value less than 5×10^−2^ but greater than or equal to 5×10^−8^.

In total (cis+trans), we identified 274 independent study-wide significant signals for 184 CSF proteins, 127 independent signals for 100 plasma proteins and 32 independent signals for 27 brain proteins (**Fig. 1, 2, Table S3, S4 and S5**). The number of significant pQTL was proportional to the sample size, rather than the number of proteins. Of the 274 study-wide significant associations in CSF, 82% were cis associations and 18% were trans associations. In plasma, 76% were cis associations and 24% were trans associations. Lastly, in brain, 94% were cis associations and 6% were trans associations (**Fig. 1b)**. The lower number of trans-pQTL in brain is likely due to the smaller sample size, and we predict that a similar proportion of cis/trans signals will be found in brain when larger studies are performed.

**Fig 2.**
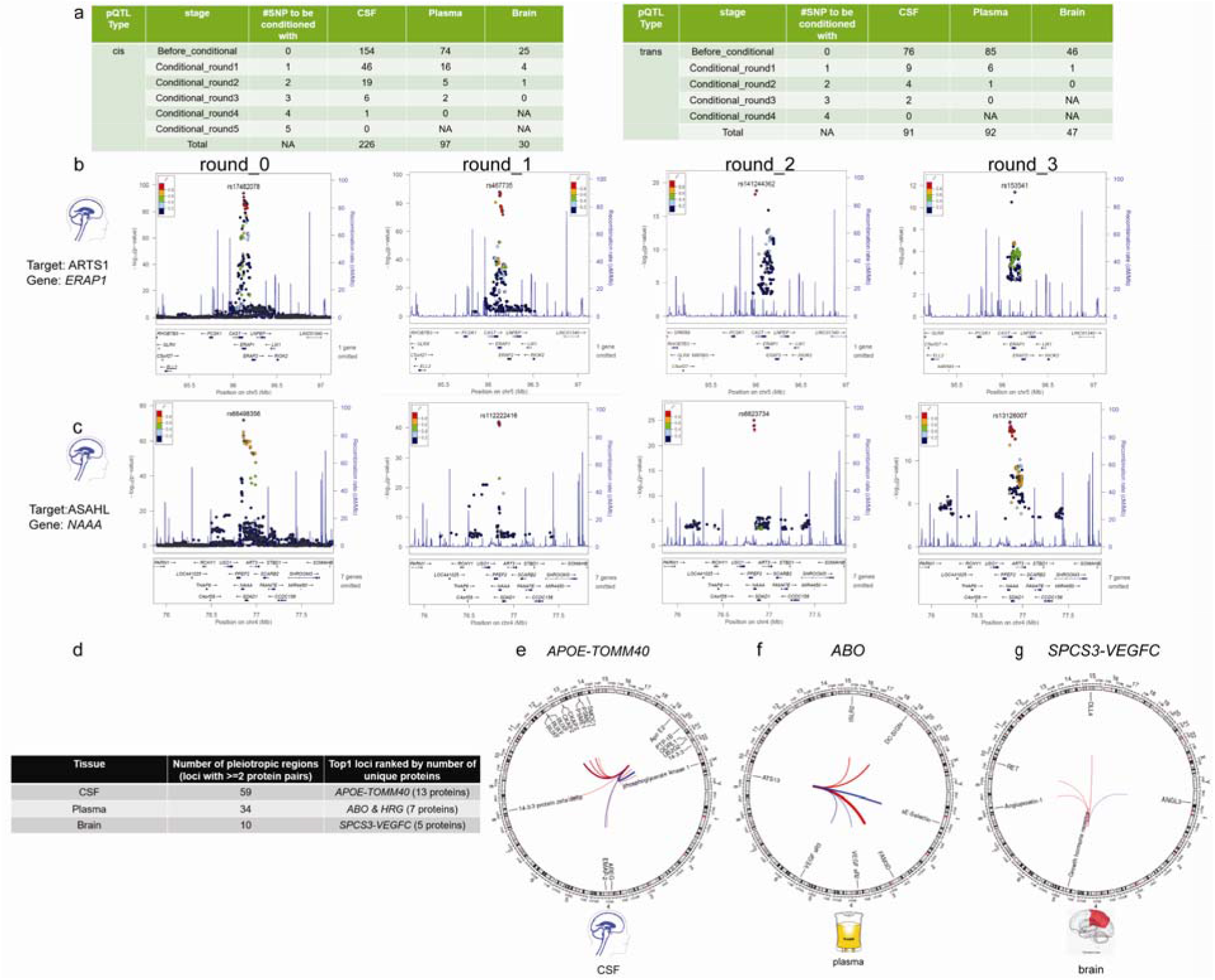
Overview of the independent loci from the conditional analyses and Identification of pleiotropic regions. (**a**) Tables of conditionally independent pQTL (cis and trans) locally (2 Mb window) after each round for each tissue. Before conditional, no SNPs were used as a covariate given one locus. For round-1 conditioning, the top SNP from before-conditioning stage given the same locus was used as an additional covariate in the default model. For round-2 conditioning, the top SNP from before-conditioning stage and top SNP from round-1 stage was used as an additional covariate in the default model. Both SNPs were within the same locus. For each round we added the previous independent top hits from the prior rounds until no variants passed genome-wide significance threshold given the same locus. **(b)** Regional association plots of the ERAP1 region associated with CSF ARTS1 protein: (round_0) before conditional analyses, centered on rs17482078; (round_1) after conditioning on the prior top SNP (rs17482078, centered on rs467735; (round_2) after conditioning on the prior top SNPs (rs17482078 and rs467735, centered on rs141244362; (round_3) after conditioning on the prior top SNPs (rs17482078 and rs467735 and rs141244362, centered on rs153541. No genome-wide significant SNPs was observed in round_4 after conditioning on all prior top SNPs. **(c)** Regional association plots of the NAAA region associated with CSF ASAHL protein: (round_0) before conditional analyses, centered on rs66498356; (round_1) after conditioning on the prior top SNP (rs66498356, centered on rs112222416; (round_2) after conditioning on the prior top SNPs (rs66498356 and rs112222416, centered on rs6823734; (round_3) after conditioning on the prior top SNPs (rs66498356 and rs112222416and rs6823734, centered on rs13126007. No genome-wide significant SNP was observed in round_4 after conditioning on all prior top SNPs. The SNPs for each regional plot are denoted as a purple diamond. Each dot represents individual SNPs, and dot colors in the regional plots represent linkage disequilibrium with the named SNP at the center. Blue vertical lines in the regional plots show recombination rate as marked on the right-hand Y-axis. **(d)** Table of all pleiotropic regions with each tissue given genome-wide significance threshold for both cis and trans-pQTL and the name of top-1 locus ranked by number of unique proteins. **(e)** Circos plot on top-1 locus (mapped to APOE-TOMM40) associated with 13 unique CSF proteins. **(f)** Circos plot on top-1 locus (mapped to ABO or HRG) associated with 7 unique plasma proteins. **(g)** Circos plot on top-1 locus (mapped to SPCS3-VEGFC) associated with 5 unique brain proteins. Outermost numbers denote chromosomes. Lines link the genomic location of this locus with genes encoding significantly associated proteins. Associations denote genome-wide significance. Line thickness is proportional to effect size of linear regression (red, positive; blue, negative).

Previous studies discovered that most eQTL are non-coding variants leading to the hypothesis that most of the eQTL effects act through modulation of transcription factor binding or chromatin structure^14^, but it is not clear if this is the case for pQTL. For this reason, we performed bioinformatic functional annotation and statistical analyses to determine if pQTL are enriched in specific regions such as untranslated regions (UTRs), downstream or upstream of the gene, splice sites, Non-coding RNA (ncRNA) splice sites, ncRNA_introns, ncRNA_exons, introns, intergenic regions, or exons. We found that the strength of the association (effect size or beta) for cis signals decreased with distance from the transcription start site (**Extended Fig**.**3a**), similar to what has been previously reported for cis-eQTL^14^. This effect was found in all three tissues, suggesting that this is a common biological event. There was an inverse relationship between absolute value of the effect size (beta) and MAF (**Extended Fig**.**3b**), consistent with previous protein level genome-wide association studies (p-GWAS;^7,15^). However, both cis and trans pQTL were strongly enriched for exonic variants (Odds Ratio = 3.71, 5.25, 4.19 for CSF, plasma, and brain respectively; **Extended Fig**.**3c)**. In 42-53% of the cis effects (95 out of 226 in CSF, 44 out 97 in plasma, 16 out of 30 in brain), the association can be explained by a coding variant whereas in eQTL only 2-5% ^5^ of the signals are located in coding regions, indicating that pQTL are significantly enriched for coding variants. These results suggest that are additional regulatory mechanisms (including post-transcriptional changes) for protein levels that may not be implicated for mRNA levels.

The enrichment of coding variants for pQTL was not only in cis but, but also in trans (**Extended Fig**.**3, Extended Fig**.**6, Table S3, S4, S5**), suggesting that protein levels are more likely to be regulated post-transcriptionally than by regulating mRNA levels. In multiple cases, the most significant signal was a coding variant in the gene that affects protein cleavage or secretion (cis-signal; as in the case of IL6R or YKL-40; **Extended Fig**.**3c, Table S3**), or a coding variant in the receptor of the protein that is likely to modify protein-receptor binding (trans-signal: as in the case of variants in the *APOC4* gene region that are associated with the BAFF Receptor; **Extended Fig**.**3c, Table S3**). In line with the hypothesis that coding variants have a higher effect size and that pQTL are enriched for coding variants, we found that pQTL explain a large proportion of the variation in protein levels. The median variation in protein levels explained by pQTL was 9% to 14.9% (interquartile range: 13.2% to 15%; **Extended Fig**.**3d**). However, there are some extreme cases in which the top pQTL explains more than half of the variability in protein levels, such as in the case of rs2075803 (p.K100E) which explains 81% of the CSF Siglec-9, rs5498 (p.K469E) which explains 74.4% of plasma sICAM-1, and rs5498 (p.K469E) which explains 67.4% of brain PPAC (**Extended Fig**.**3d, Table S3, S4, S5**). The CSF Siglec-9, plasma sICAM-1, and brain PPAC have been replicated in other studies^13,15–17^ using a different proteomic approach indicating that these findings are not a platform driven finding.

To replicate our pQTL findings, we accessed, processed and analyzed several publicly-available datasets. We also performed meta-analyses and cross-tissue replication. For CSF, we leveraged two studies in which similar proteomic and genetic data were available: Sasayama et al. generated aptamer-based proteomic and GWAS data from 132 samples of Japanese origin^11^ and the Parkinson’s Progression Markers Initiative (n=131; released December 2019, PPMI19 hereafter), also had proteomics and genetic data available. We found that 51 pQTL (49 cis and 2 trans) were genome-wide significant and in the same direction in those studies. We also performed a meta-analysis of the Sasayama and the PPMI studies (**Table S6**) to identify additional genome-wide and nominal associations. We found that 153 of our study-wide significant signals had genome-wide significance in PPMI+Sasayama meta-analysis and 27 additional pQTL showed at least a nominal association in the same direction in the Sasayama+PPMI meta-analyses. We also identified an additional 16 pQTL that have been reported to be pQTL in other tissues (plasma in AddNeuroMed ^18^, INTERVAL^7^, KORA^8^, SCALLOP^9^, and a mass spectrometry based pQTL dataset^13^, and in our plasma or brain pQTL data). We were unable to test for replication of five pQTL as protein levels were not available in other studies. In summary, we were able to replicate more than 90.1% of the CSF pQTL, which is higher than previous studies^7^. Twenty two pQTL are still pending replication, as current studies with smaller sample sizes do not provide enough statistical power. However, based on our validation with plasma and brain pQTL, a good proportion are likely real. This is supported by the fact that we have been able to replicate 96.8% and 96.9% of the plasma and brain pQTL (see below).

For replication of the plasma pQTL we used the AddNeuroMed (n=343), INTERVAL (n=3,301), KORA (n=1335), and SCALLOP (n=3394) studies. We were able to replicate 96.8% of our 127 pQTL. We were not able to test another two, as they were not measured in those studies. For brain, there were no published studies using the same aptamer-based proteomic method. However we matched our proteins with a mass spectrometry based pQTL dataset^13^. We were able to replicate five signals at genome-wide significance, eight signals at a nominal association, and 17 pQTL that showed at least a nominal association in brain or CSF. Only one pQTL was not replicated.

To increase the statistical power and identify additional genome-wide significant pQTL, we performed meta-analyses that included all the CSF cohorts as well as multi-tissue analyses. We first performed a CSF mega meta-analysis including the 596 common proteins shared among our study, PPMI and the Sasayama study (**Extended Fig**.**4c**,**d**). The additional sample size identified 425 pQTL for 310 proteins, compared to the 250 pQTL for 185 proteins identified by our CSF cohort. This represents an almost two-fold increase in the pQTL signals by increasing the sample size just 25%, suggesting that many other pQTL will be identified with a larger sample size. We observed a similar increased in the number of pQTL when performing a multi-tissue meta-analysis. For these analyses, we included 342 proteins that passed QC in our three tissue types as well as the PPMI and Sasayama study, and found 253 pQTL compared to the 139 that were found in our CSF sample (**Extended Fig**.**4c**,**d**).

Because our study includes cognitively normal elderly individual and Alzheimer disease cases, we performed additional analyses to determine if any of the pQTL are disease-specific. We included disease status as a covariate, and performed case-only and control-only analyses and compared the effect sizes (betas) of the genome-wide pQTL of the initial analyses with the beta of these analyses. We found an extremely high correlation (Pearson’s r>0.98, **Extended Fig**.**5, Table S7, S8, S9**), indicating that the association of the genetic variants with protein levels does not depend on disease status.

## Complex and pleiotropic loci

To identify complex loci where more than one independent association exists at the same region, we performed conditional analyses for the top GWAS signal. After a first round of conditional analyses, 87 CSF, 30 plasma, and 6 brain loci still had SNPs with independent and genome-wide significance (**Fig. 2a**). We performed a second round of conditional analyses including the top SNP from the initial and the first conditional analyses. In this second round of conditional analyses 32 CSF, eight plasma, and one brain loci still showed independent and genome-wide significant association (**Fig. 2a)**. We continue performing conditional analyses until no SNP passed the genome-wide threshold. We found one protein with up to five independent signals, ten with four and more than 30 with three independent signals. There were still nominal associations after the conditional analyses for a large proportion of the loci. This indicates that we may be underestimating the number of independent signals for these loci, and highlights the complex mechanisms that regular protein levels.

As an example of these complex regions: the main signal for CSF ARTS1 (**Fig. 2b**) is driven by the nonsynonymous variant p.D575N (rs10050860; p-value=7.41×10^−86^). After conditioning on this signal, there was still a genome-wide signal tagged by rs469674 (conditional p-value=1.40×10^−88^) which is predicted to affect a transcription factor binding site (RegulonDB score =1^19^) and is associated with gene expression (GTEx; multi-tissue p-value=0). The third independent signal in this region (rs27895; conditional p-value=2.17×10^−13^) is driven by a nonsynonymous variant (p.G346D). Similarly, the main signal for CSF ASAHL is driven by the nonsynonymous variant p.V151I (rs4859571; p-value=4.79×10^−63^) and the second independent signal by rs7688400 (conditional p-value=1.82×10^−42^) which affects gene expression (RegulonDB Score=2; GTEx p-value:3.25×10^−103^). The third independent signal is tagged by the nonsynonymous variant p.F334L (rs6823734, conditional p-value=1.04×10^−25^), and the fourth independent signal is tagged by another coding variant (**Fig. 2c**). All together, these results indicate that proteins are highly regulated and include several independent mechanisms, even at the same locus. These mechanisms may affect protein expression levels by affecting cleavage, cell secretion, receptor binding, or clearance (nonsynonymous variants) and others by regulating gene expression (non-coding variants).

We found that there are some loci that regulate the levels of not only one protein, but up to 13 different proteins as in the case of genetic variants in the *APOE* region. Genetic variants in the *APOE* gene region are associated with 13 different CSF proteins, including PTP-1B, Apo E2, SMOC1, EMAP-2, AREG, 14-3-3, phosphoglycerate kinase 1, CKAP2, RUXF, 14-3-3 protein_zeta/delta, PSME1, UB2G2 and QORL1 (**Fig. 2d**,**e, Extended Fig**.**6a, Table S10**). These proteins are located on different chromosomes indicating that this is not just a cis-regulation, where these genes present similar mRNA regulatory mechanism because they are located on the same chromosomal region. Genetic variants in *APOE* are the strongest risk factors for Alzheimer disease and Lewy body dementia. Several studies have found that the 14-3-3 protein is a marker of non-specific neuronal death^20,21^, and our results supports 14-3-3 protein_zeta/delta as a potential biomarker for Alzheimer disease and other neurodegenerative diseases. For CSF, we found 59 pleotropic regions where the same locus was associated with two or more proteins. In plasma, we replicated the known pleiotropy of the *ABO* locus for seven different proteins including E-Selectin (**Fig. 2f, Extended Fig**.**6b, Table S11**), which was implicated in stroke risk by recent studies using Mendelian Randomization approaches^22^. Further studies are needed to establish how these, and other genetic variants are associated with more than one protein. For example, genetic variants in the SPCS3-VEGFC region regulates brain levels of five different proteins: Angiopoietin-1, Growth hormone receptor, RET, ANGL3, and DLL4 (**Fig. 2g, Extended Fig**.**6c, Table S12**). Similarly, F5 regulates CSF Coagulation Factor V, CAMK1, P-Selectin and L-Selectin. An additional 32 pleotropic regions in plasma and nine in brain tissue were found.

Many of the published studies on eQTL and pQTL have not identified pleotropic regions because they have focused on cis-associations^13,23–25^. Our results indicate that protein expression is regulated at multiple levels and in coordination with other independent proteins, which are likely part of the same signaling pathway. In order to understand the biological process of health and disease, it is important to identify which proteins are regulated by the same genetic factors. This study identifies tissue-specific pleiotropic effects, highlighting the complex mechanisms that regulate protein levels. Identification of additional tissue-specific cis, trans and pleotropic regions will identify novel pathways relevant to pathogenesis.

## Multi-tissue genetic architecture of protein levels

Our unique study design, which includes high-throughput protein levels in multiple tissues linked to genetic data, enabled us to investigate the overlap of the genetic architecture of protein levels across tissues. We compared the overlap of study-wide significant cis and trans-pQTL between CSF, plasma and brain, while focusing on the proteins (n=411) that passed QC in all three tissues (**Fig. 3a, Table S13**). We observed that cis-pQTL are more likely to be shared across tissues than trans-eQTL. When using the standard genome-wide significance cutoff as a threshold, we found that 11.8% (21 of 199) of cis-pQTL loci were shared between the three tissues, and 48% (86/178) of cis-pQTL are found in two or more tissues. Although cis-pQTL are often shared between tissues, more than half of the pQTL (92/178) were tissue-specific. Only one trans-pQTL was shared across all tissues and only 14.0% (19 of 135 trans) are shared between two or more tissues. Plasma shared more pQTL with CSF than brain (**Fig. 3b**,**c**; in cis: 72 vs 24; and trans:19 vs 1). CSF seems to capture the overall genetic architecture of brain protein levels better than plasma, as 91% (32/35) of the brain cis-pQTL are found in CSF compared to the 68% (24/35) captured by plasma.

**Fig 3.**
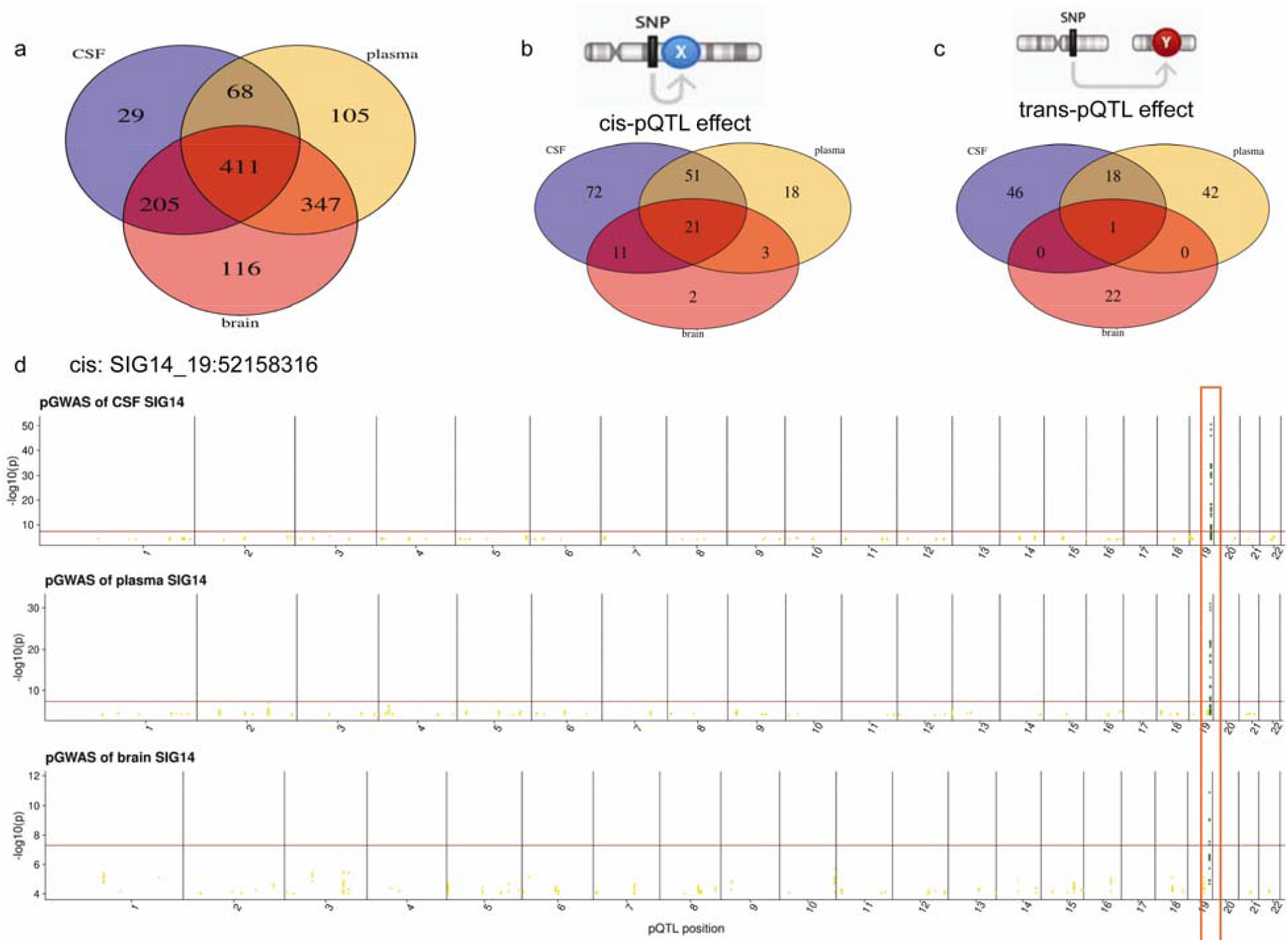
Summary of the tissue-specificity analyses. (**a**) Venn diagrams of proteins passing QC across all three tissues. (**b**) Venn diagrams of all cis-pQTL across all three tissues given P< 5×10^− 8^ threshold. (**c**) Venn diagrams of all trans-pQTL across all three tissues given genome-wide significance threshold. (**d**) Manhattan plots of the ISG14-chr19:52158316 within each tissue as an example of three-tissue-shared cis-pQTL.

As our sample size was not consistent across tissues and CSF had a higher number of samples, these results could be biased by differential statistical power. Therefore, we performed similar analyses with more permissive p-values. We contemplated two complementary scenarios: a pQTL is shared across tissues if it is genome-wide significant for one tissue and has a p-value of <0.05 (scenario 1; **Extended Fig**.**7a, Table S14**) or p<0.001 (scenario 2; **Extended Fig**.**7b, Table S15**). These two scenarios led to similar results as those performed with the genome-wide threshold. Between 22-32% of the cis-pQTL were found in all the tissues compared to 2-4% of the trans-pQTL, and CSF was able to capture most of the cis and trans brain pQTL. We performed similar comparisons by comparing our CSF and brain pQTL with the plasma pQTL from the INTERVAL study that includes 3,301 samples to confirm that our findings were not an artifact of the sample size. For these comparisons we focused on 616 proteins that passed QC in CSF and brain from this study, and plasma from INTERVAL (**Extended Fig**.**8a-d**). These analyses replicated our findings and indicate cis-pQTL are more likely to be shared across tissues than trans-pQTL. As expected, CSF captures the overall genetic architecture of brain protein levels better than plasma, but plasma still captures more than half of the observed brain pQTL, suggesting that plasma may still be an informative tissue to study brain-related disorders, such as Alzheimer disease.

## Multi-tissue pQTL identify functional genes and provide a biological context for GWAS findings

eQTL mapping and colocalization analyses have been instrumental in identifying the functional genes in genetic studies of complex traits^5,26^. However, it is known that changes in transcript levels do not necessarily translate to changes in protein levels. In order to resolve all GWAS signals, it is vital to determine the overlap between eQTL and pQTL. We analyzed the overlap of our cis-pQTL with the latest release GTEx v8 cis-eQTL across all 50 tissues (**Table 2 & Table S16**).

**Table 2.**
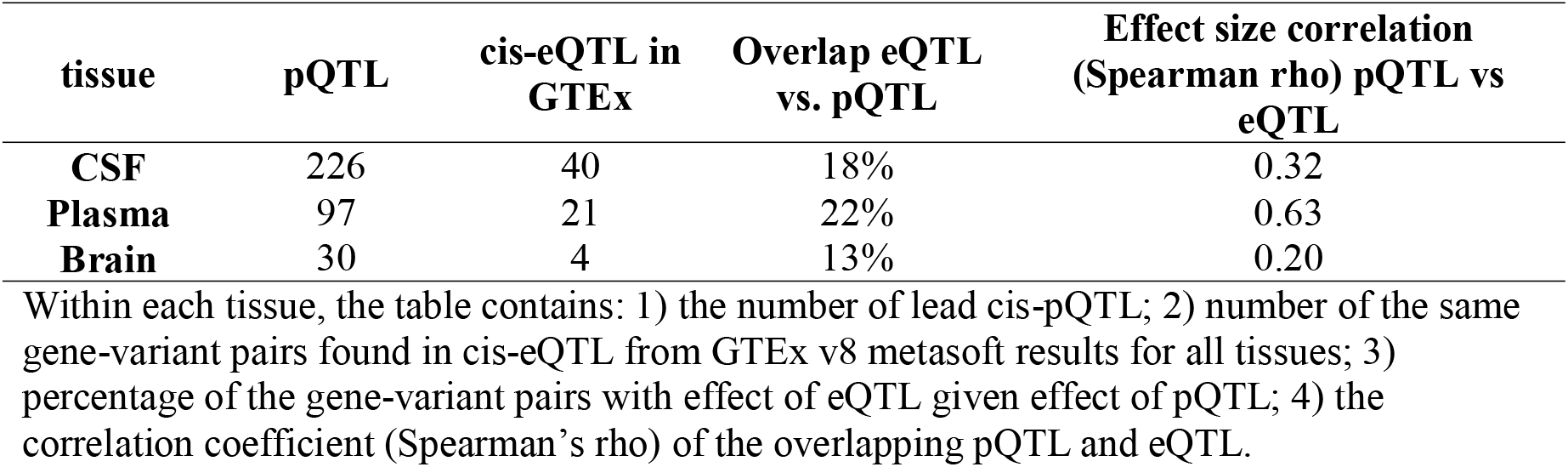
Overlap of cis-eQTL from GTEx and cis-pQTL from this study.

| tissue | pQTL | cis-eQTL in GTEx | Overlap eQTL vs. pQTL | Effect size correlation (Spearman rho) pQTL vs eQTL |
| --- | --- | --- | --- | --- |
| **CSF** | 226 | 40 | 18% | 0.32 |
| **Plasma** | 97 | 21 | 22% | 0.63 |
| **Brain** | 30 | 4 | 13% | 0.20 |

Within each tissue, the table contains: 1) the number of lead cis-pQTL; 2) number of the same gene-variant pairs found in cis-eQTL from GTEx v8 metasoft results for all tissues; 3) percentage of the gene-variant pairs with effect of eQTL given effect of pQTL; 4) the correlation coefficient (Spearman's rho) of the overlapping pQTL and eQTL.

Only 13-22% of the cis-pQTL were also cis-eQTL for the same gene from the GTEx multi-tissue analysis. More specifically, in brain, only 13% of the pQTL were found in GTEx, which is similar to the Robins et al. (2019) study^13^ in which the authors used mass-spectrometry based proteomics to measure 7901 proteins in 144 samples, and found an overlap in 16% of proteins with eQTL from GTEx. This may be also explained by the fact that as the number of brain samples in GTEx is only 323 compared to the 380 samples included in this study, which represent the largest brain pQTL performed so far. In plasma, only 22% of our pQTL were also eQTL, similar to other previous studies^7^. No previous studies have analyzed the overlap between pQTL and eQTL in CSF yet. Based on our results only 18% of the pQTL have been reported as eQTL in GTEx.

We also analyzed the correlation in the for overlapping pQTL and eQTL. We found that the correlation of the QTL effect size in blood (Spearman r=0.63) was higher than in CSF (Spearman r=0.32) or brain (Spearman r=0.20). This indicates that a large proportion (37 to 80%) of the pQTL cannot be inferred directly from eQTL, especially in brain, and therefore additional multi-tissue proteomic analyses in larger datasets are needed in order to map the additional GWAS signals. As most of the eQTL analyses are only focused on cis-eQTL mapping, we were not able to analyze the overlap for the trans-eQTL.

It is critical to perform trans-pQTL analyses, because trans-pQTL analysis can identify novel protein-protein interactions or proteins that are part of the same pathway. For example, variants in the *HLA-DQA1* gene region are known to be associated with Alzheimer Disease risk^3^ and here we found that they are also associated SLAM family member 5 (SLAF5), and CD33 protein levels. *CD33* is also a known GWAS locus for Alzheimer Disease risk, and both CD33 and SLAF5 are microglia-specific proteins^27^. Recent studies, especially the identification of the association of *TREM2* with Alzheimer disease risk^28,29^, have highlighted the importance of microglia in Alzheimer disease. TREM2 acts downstream of CD33 in modulating the microglial pathology in Alzheimer disease^30,31^. Similarly, in a recent study we found a trans-pQTL for CSF TREM2 levels in the MS4A4A gene^32^. Although the two genes have been implicated in Alzheimer Disease, Deming et al. (2019) demonstrate for the first time that that MS4A4A interacts with TREM2, and that it is possible to modulate TREM2 levels by targeting MS4A4A levels. This study also provided a biological context for the original association of MS4A4A. MS4A4A modifies Alzheimer disease risk by regulating TREM2 levels. Mendelian Randomization studies also demonstrated that higher TREM2 levels are protective, and there are currently clinical trials that targets TREM2 by using activating antibodies or MS4A4A as a potential therapy for Alzheimer disease^32^. Here for the first time we are putting another two genes, HLA-DQA1 and SLAF5, in the same pathway as CD33, TREM2 and MS4A4A. These genes are likely to affect microglia activity and innate immune response This study goes beyond Alzheimer disease, and the data generated here can be leveraged for other traits as well. Multiple groups^7,9^ have also identified a trans-pQTL for the ABO locus in E-selectin that has been demonstrated to be the gene driving the association for stroke risk and may be a useful potential biomarker for stroke risk. These and other similar findings are a clear example of how pQTL studies can help identify the functional genes underlying GWAS signals, as well as add biological context to the GWAS loci.

## Mendelian Randomization to identify novel biomarkers and drugs for neurodegenerative diseases

We and others^10,33^ have demonstrated that by combining pQTL summary statistics with Mendelian Randomization (MR) analyses it is possible to identify novel biomarkers and causal proteins for complex traits. The advantage of this approach is that it is possible to perform large-scale unbiased biomarker discovery for multiple complex traits, without the need for expensive studies. To infer the causality between proteins and complex traits, we performed MR analysis by using pQTL as instrumental variables. We performed MR for Alzheimer disease risk, onset and progression, Parkinson disease, Frontotemporal dementia, Amyotrophic lateral sclerosis and stroke risk (**Table S17**). We found three potential biomarkers and proteins involved in Alzheimer disease risk in CSF, and 13 in plasma (**Table 3, Fig. 4, Table S18**). Some of these proteins were regulated by known GWAS hits, but others were novel. For example, using AD risk as the outcome, Siglec-3 protein (encoded by *CD33*), a known regulator for Alzheimer disease risk^3^, was found significantly associated with Alzheimer disease risk in plasma and CSF. Higher plasma Siglec-3 (CD33) was associated with increased Alzheimer disease risk.

**Table 3.** Number of significant protein-trait associations from Mendelian Randomization analyses.

| **Outcome** | **CSF** | **Plasma** | **Brain** |
| --- | --- | --- | --- |
| **AD risk** | 3 | 13 | 7 |
| **AD progression** | 18 | 25 | 6 |
| **AD Age at Onset** | 8 | 2 | 20 |
| **PD risk** | 13 | 15 | 35 |
| **ALS risk** | 1 | 4 | 3 |
| **FTD risk** | 1 | 8 | 5 |
| **Stroke risk** | 7 | 8 | 24 |

Within each tissue, the table contains the number of significant proteins (FDR < 5/21) and associations with the seven neurological traits: 1) AD risk<sup>3</sup>; 2) AD progression<sup>63</sup>; 3) AD Age At Onset<sup>64</sup>; 4) PD risk<sup>36</sup>; 5) ALS risk<sup>65</sup>; 6) FTD risk<sup>66</sup>; 7) Stroke risk<sup>67</sup>.

**Fig 4.**
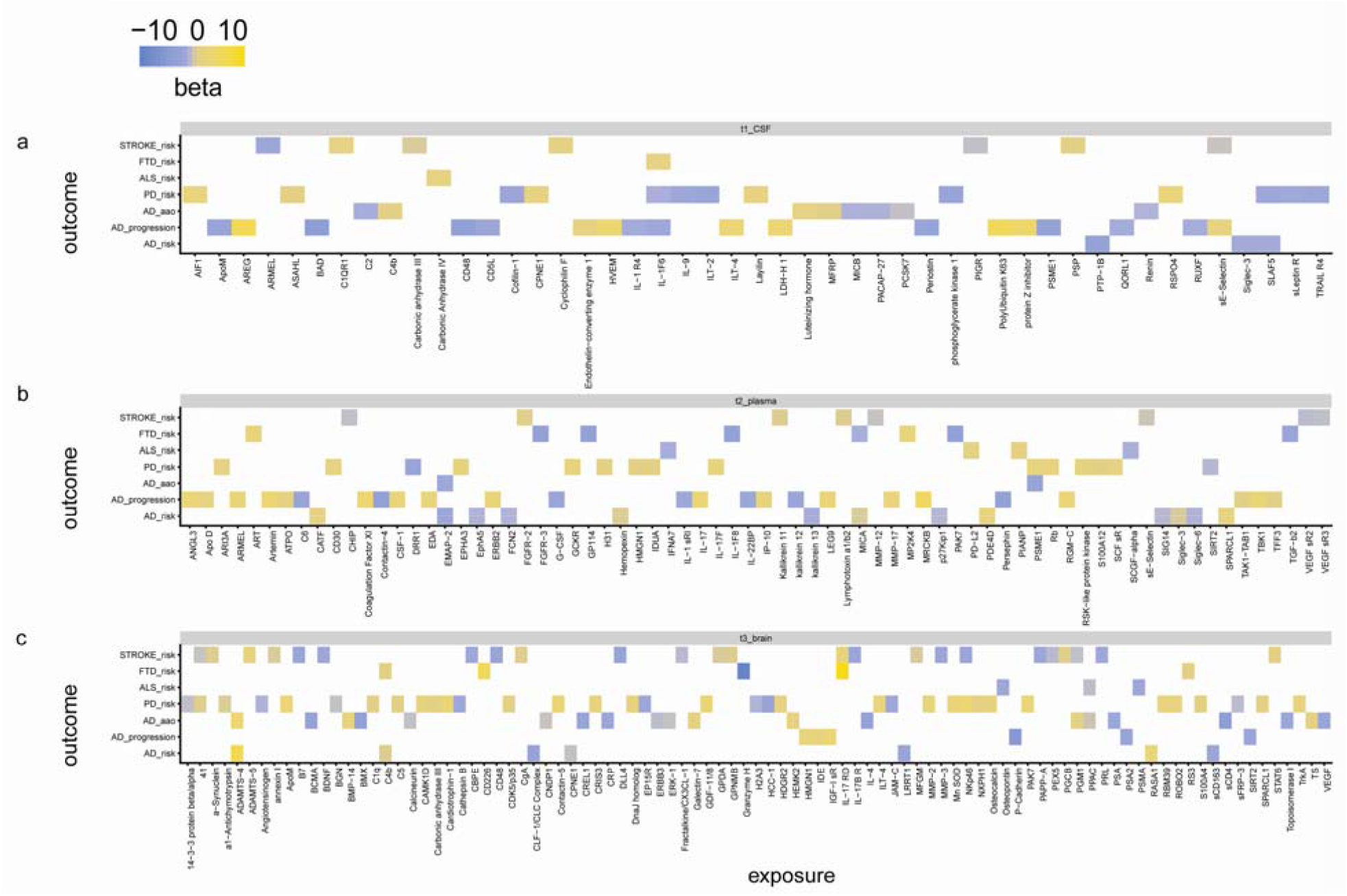
Mendelian randomization prioritized proteins in the causal relationship with seven neurological traits. Mendelian Randomization (MR) results is calculated using the “TwoSampleMR” R package ^62^, and the effects for each protein-disease pair are visualized using Heatmap of MR inference of (**a**) CSF, (**b**) plasma, and (**c**) brain protein effect on seven neurological-related traits. The p-value threshold for significance is 0.05 after multiple test correction. The color represents whether the effect size is positive (yellow) or negative (blue). Alzheimer disease (AD); Parkinson disease (PD); Amyotrophic lateral sclerosis (ALS); Frontotemporal dementia (FTD). Stroke is the general risk, not a specific subset of the stroke. The summary statistics are curated from published datasets (Table S16 for details).

The *TMEM175/GAK/DGKQ/CPLX1/IDUA* locus is the third most significant locus in the large metaLJanalyses of Parkinson disease risk GWASs^34–36^. This multi-gene locus contains more than one independent signal associated with PD^36^. However, it is unclear which gene or genes are responsible for this association. Several recent papers suggest that coding variants in *TMEM175*^37^, eQTL affecting *GAK*^36^, *IDUA* and *DGKQ*^38^ or *CPLX1*^39^ could be the drivers of this association. We found that the SNPs in this locus were associated with IDUA protein levels as well as Parkinson’s disease risk (rs35220088; p= 2.47×10^−6^ and 3.52×10^−9^, respectively) and our MR analyses indicate that *IDUA* is a functional gene in this locus. (MR FDR-corrected p-value: 9.37×10^−6^; **Table S17**). The *IDUA* gene encodes alpha-L-iduronidase, which degrades glycosaminoglycans (GAGs) in the lysosome. Mutations in the *IDUA* gene cause mucopolysaccharidosis type I (MPS I), a lysosomal storage disease^40^. These findings also highlight the power of pQTL to help in determining the causal gene in a multi-loci genomic region associated with disease risk.

E-Selectin protein is a known stroke biomarker^22^, and our MR analyses indicate that this protein is not only a biomarker but also is part of the cascade of pathogenic events that leads to disease (**Table S18**). Some proteins were inferred to be functional across more than one neurodegenerative disease within each tissue (**Table S18**). Among CSF proteins, IL-1F6 was shown in the MR analyses to be associated with Alzheimer disease progression, Parkinson’s Disease risk and Frontotemporal Dementia risk. SLAF5 was linked to both Alzheimer and Parkinson’s Disease risk. Among plasma proteins, MICA was involved in the causal pathways of both Alzheimer disease risk and Frontotemporal Dementia risk and PSME1 was involved in the causal pathways of both Alzheimer disease age at onset and Parkinson’s Disease risk. Among brain proteins, IL-17 RD was involved in causal pathways of both Frontotemporal Dementia risk and stroke risk, and PPAC was involved in the causal pathways of both Alzheimer disease age at onset and Amyotrophic lateral sclerosis risk.

Our study and those like it are not only useful for identifying pQTL that will help to resolve GWAS locus, but also for identifying novel biomarkers and causal pathways using MR analyses. We integrated the pQTL data, MR analyses and databases for approved and experimental drug targets to identify potential drugs that could be repurposed for diseases caused by complex traits. It is also known that compounds aiming to target proteins supported by genetic and genomic data are more likely to work than those without genetic support. We found that 44.5% (n=130), 37% (n=67) and 49.3% (n=39) of the CSF, plasma and brain proteins with pQTL colocalize with drug targets (**Table S19**).

Our results identify multiple potential drug repurposing opportunities. For example, we identified a very strong cis-pQTL (p= 3.4×10^−111^, and 8.3×10^−84^ in CSF and plasma respectively) for Siglec-3, which is encoded by CD33 and is a known GWAS locus for Alzheimer disease risk. AVE9633 is an anti-CD33 antibody used in acute myeloid leukemia, however our studies indicate that CD33 could be also be a target for Alzheimer disease. In fact, there are already clinical trials that are using antibodies targeting CD33 as a potential therapy for Alzheimer disease^41^. Similarly, our results suggest that Acetazolamide as a potential drug for amyotrophic lateral sclerosis. Our genetic analyses found a strong pQTL for carbonic anhydrase IV, which is a target for Acetazolamide. Several studies have reported that the expression of Carbonic Anhydrase IV is altered in the motor neurons of Amyotrophic lateral sclerosis patients^42,43^. Acetazolamide is a carbonic anhydrase inhibitor^44^, used to treat glaucoma, epilepsy and altitude sickness^45^. Acetazolamide has been associated with worse outcomes in Amyotrophic lateral sclerosis, supporting the idea that targeting carbonic anhydrase IV can modify disease course of Amyotrophic lateral sclerosis. However, our results indicate that higher activity levels of carbonic anhydrase IV would be associated with better Amyotrophic lateral sclerosis outcomes. Finally, our analyses nominated *IDUA* as the functional gene for the Parkinson’s disease TMEM175/GAK/DGKQ/CPLX1/IDUA locus, but also indicate that *IDUA* could be a potential drug target. These data indicate that *IDUA* can be targeted with chondroitin sulfate. IDUA is required for the lysosomal degradation of glycosaminoglycans, dermatan sulfate and heparan sulfate (HS). HS significantly stimulates the formation of α-Synuclein fibrils *in vitro*^46^. HS also mediates macropinocytotic uptake of α-Synuclein^47^. Pharmacological inhibition of HS binding of α-Synuclein and genetic reduction of HS synthesis facilitates the clearance of pathogenic proteins and reduce their aggregation^47^. Additional potential drugs targeting proteins with strong pQTL can be found in **Table S19**. Overall, our data and analyses provide new evidence for potential therapeutic targets by linking genetic factors to disease via specific proteins.

## Discussion

In this study we generated a detailed map of multi-tissue pQTL that will be instrumental in understanding the tissue-specific genetic architecture of protein levels. pQTL were enriched for coding variants compared to eQTL. By leveraging this data, we have been able to resolve some of the GWAS loci, identify additional proteins implicated in disease pathogenesis and identify novel biomarkers for complex traits. If we want to understand the genetic architecture of complex traits, we first need to understand the genetic architecture of protein levels. Until now, gene expression has been the main approach to solve some of the GWAS loci. However, here we demonstrate that a large proportion (>78%) of the pQTL are likely driven by post-transcriptional and post-translational effects, and therefore cannot be found by eQTL analyses. However, multi-tissue pQTL mapping has been constrained by the limited availability of large-scale proteomic analyses in several tissues, a bottleneck remedied by this study. Here we present the largest brain and CSF pQTL analyses performed so far, the first neurological-relevant multi-tissue pQTL study and a unique resource to leverage multi-tissue pQTL to understand complex traits. This data can be leveraged to perform MR analysis on other CNS pathologies for which there is good GWAS data such as schizophrenia or bipolar disorders, as well as cognition or measures of brain function. All the results from this study can be interactively accessed through the Online Neurodegenerative Trait Integrative Multi-Omics Explorer (ONTIME; https://ontime.wustl.edu/).

## Methods

### Aptamer-based proteomics data sample collection

This study comprises about 1237 participants from Washington University. Samples from participants were of three tissue types: a) CSF; b) plasma; c) brain (parietal lobe cortex). CSF samples were collected in the morning after an overnight fast, processed, and stored at −80 °C. Plasma samples were collected in the morning after an overnight fast, immediately centrifuged, and stored at − 80°C. Brain tissues (∼500mg) were collected from the fresh frozen human parietal lobes.

For CSF tissue, there were 971 unique participants and 1300 samples (329 participants provided two samples, one baseline and one longitudinal) in total, including 275 AD cases, 1020 cognitively normal controls and five with unknown status. The age is normally distributed with a mean of 69.4 years and standard deviation of 9.3 years. 53% of the samples are from women. For plasma tissue, there are 636 unique participants and 648 samples in total, including 234 AD individuals, 409 cognitively normal controls and five with unknown status. The age is normally distributed with a mean of 69.8 years and standard deviation of 9.4 years. 54% of the samples are from women. For brain tissue, there are 458 unique participants and 459 samples in total, including 345 cases, 12 cognitively normal controls and 102 with unknown or other status (e.g. FTD, other neurological diseases). The age is normally distributed with a mean of 83.3 years and standard deviation of 10 years. 57% of the samples are from women. The donor overlap across three tissues are shown as a Venn diagram in **Extended Fig. 2b**: 9 donors were shared across all three tissues; 481 donors were shared by both CSF and plasma; 29 donors were shared by plasma and brain; 481, 117 and 420 were exclusively for CSF, plasma, and brain tissues respectively. The Venn Diagram was drawn using VennDiagram^48^ R package. These recruited participants were evaluated by the clinical personnel from Washington University. The AD severity was determined by the Clinical Dementia Rating (CDR)^49^. The Institutional Review Boards of Washington University School of Medicine in St. Louis approved the study, and research was carried out in accordance with the approved protocols.

### Proteomic data QC process

We used a multiplexed, aptamer-based approach^50^, to measure the relative concentrations (relative fluorescent units, RFU) of proteins from CSF, plasma and brain tissues, assayed using 1,305 modified aptamers in total. The assay covers a 10^8^ dynamic range, and measures all three major categories: secreted, membrane and intracellular proteins. The proteins cover a wide range of molecular functions, such as protein binding and the MAPK cascade. The coverage of proteins on the platform has taken into account proteins known to be relevant to human disease, including Alzheimer diseases ^51^, cardiovascular diseases^52^, thus has been widely used for biomarker discovery.

Aliquots of 150 μl of tissue samples were sent to Genome Technology Access Center at Washington University in St. Louis for protein measurement. Assay details have been previously described by Gold and colleagues ^50^. In brief, modified single-stranded DNA aptamers are used to bind to specific protein targets that are then quantified by a DNA microarray. Protein concentrations are quantified as RFU.

Quality control (QC) was performed at the sample and aptamer levels using control aptamers (positive and negative controls) and calibrator samples. At the sample level, hybridization controls on each plate were used to correct for systematic variability in hybridization. The median signal over all aptamers was used to correct for within-run technical variability. This median signal was assigned to different dilution sets within each tissue: For CSF samples, a 20% dilution rate was used. For plasma samples, three different dilution sets (40%, 1% and 0.005%) were used. For brain samples, a 20% dilution rate was used.

To QC the proteomics datasets (**Extended Fig**.**1a-d**), the protein/analyte outliers were first removed by applying four criteria as below: 1) Minimum detection (LOD) filtering. Limit of detection (LOD) was defined as the summation of average expression level of the new NP-buffer (used as dilution buffer of CSF samples since plate-42) and K fold of standard deviation (K = 2). If the analyte for a given sample was less than LOD, this sample was an outlier. Collectively, if the number of outliers given an analyte was less than 15% of total sample size, this analyte was kept. 2) Flagging analytes based on the scale factor difference. The scale factor difference was calculated as the absolute value of the maximum difference between the calibration scale factor per aptamer and the median for each of the plates run. If the value for this analyte was less than 0.5 (NOTE: SOMAlogic SQS report used 0.4), the analyte passed this criterion. 3) CV of calibrators lower than 0.15. The coefficient of variation (CV) for each aptamer was calculated by dividing the standard deviation by the mean of each calibrator at the raw protein level. If the analyte had a CV of less than 0.15, this analyte passed the CV QC. 4) IQR strategy. Outliers were identified if the subject was located outside of either end of distribution using a 1.5-fold of IQR given the log10 transformation of the protein level. Collectively, if the number of outliers of given an analyte was greater than 85% of total sample size, this analyte was filtered. Analytes were kept after passing all the criteria above for the downstream statistical analysis. 5) An orthogonal approach was used to call subject outliers based on IQR. Collectively, if the number of outliers given an analyte was greater than 85% of total number of analytes passed QC, this subject was labeled as an outlier.

Furthermore, the analytes were removed if shared by most (∼80%) of the subject outliers. After this second removal of analytes, subject outliers were called again. Outlier subjects were again removed.

Proteins were mapped to UniProt^53^ identifiers and Entrez gene symbols. Mapping to Ensembl gene IDs and genomic positions (start and end coordinates) was performed using gencode v30 liftover to hg19/GRCh37.

### Reproducibility investigation via comparisons between biological or technical replicates

To measure the reproducibility of the aptamer-based platform, we compared the replicates for the same subject given each tissue.

For plasma samples, we included 11 subjects with two measures, one as fasted and the other as non-fasted. After QC, we kept 931 proteins in 633 samples. The overall Pearson’s correlation coefficient between fasted and non-fasted samples (**Extended Fig**.**2d**) is 0.907, with a 95% confidence interval from 0.904 to 0.911 and p-value < 2.2 × 10^−16^.

For plasma samples, we included one subject with two biological replicates: one collected in 1997, the other in 2007. Both samples passed QC. The overall Pearson’s correlation coefficient between these two biological replicates (**Extended Fig**.**2e**) is 0.938, with a 95% confidence interval from 0.9299 to 0.9453 and p-value < 2.2 × 10^−16^.

For brain samples, we included one subject with two technical replicates. After QC, we kept 1079 proteins and 435 samples. Out of these 435 samples, only one replicate of the subject remained. Thus, we compared two technical replicates using the values before QC across all 1305 proteins. The overall Pearson’s correlation coefficient between these two replicates (**Extended Fig**.**2f**) is 0.976, with a 95% confidence interval from 0.9762 to 0.9812 and p-value < 2.2 × 10^−16^.

For CSF samples, we designed 329 subjects with two measures, one as baseline (LP date1) and the other as longitudinal (LP date 2). After QC, we kept 713 proteins and 1270 samples. Out of these 1270 samples, 321 subjects with two measures remained in the analysis (**Extended Fig**.**2a**,**c**). The average time difference between the two LP dates was 6.14 years, and the standard deviation was 2.98 years. The overall Pearson’s correlation coefficient (**Extended Fig**.**2c**) between two LP dates was 0.995, with a 95% confidence interval from 0.99518 to 0.99526 and p-value < 2.2 × 10^−16^.

The overall high correlations within each tissue indicated that the aptamer-based technology was highly reproducible.

### Genomic data QC process

Most of the samples with proteomic profiling were collected with genotyping data (**Extended Fig**.**1d**). For CSF, 965 out of 971 unique subjects have both genotype and proteomic data. For plasma, 633 out of 636 unique subjects have both genotype and proteomic data. For brain tissue, 450 out of 458 unique subjects have both genotype and proteomic data.

Samples were genotyped on multiple genotyping platforms from Illumina. Stringent quality thresholds were applied to the genotype data for each platform separately. SNPs were kept if they passed all of the following criteria: i) genotyping successful rate >= 98% per SNP or per individual; ii) MAF >= 0.01; iii) Hardy-Weinberg equilibrium (HWE) (p>=1 × 10^−6^). After removing low quality SNPs and individuals, genotype imputation was performed using the Impute2 program with haplotypes derived from the 1,000 Genomes Project (released June 2012). Genotype imputation was performed separately based on the genotype platform used. SNPs with an info-score quality of less than 0.3 reported by Impute2, with a MAF < 0.02 or out of HWE were removed. After Imputation and QC, the different imputed plink files were merged. A total of over 14 million (14,059,245) imputed and directly-genotyped SNPs and 1,530 individuals were used for final analyses. To adjust for population substructure (**Extended Fig**.**1d**), principal component analysis (PCA) was conducted using the PLINK1.9 (v1.90b6.4)^54^ subcommand pca. HapMap samples (CEU: Caucasian Europeans from Utah; JPT: Japanese in Tokyo; YRI: Yoruba in Ibadan, Nigeria) were included in the analyses in order to remove outliers and confirm self-reported ethnicity. Samples kept were within the CEU cluster. To identify unanticipated duplicates and cryptic relatedness using pair-wise genome-wide estimates of proportion identity by descent (IBD) (**Extended Fig**.**1d**), we used the subcommand IBD from PLINK1.9 (v1.90b6.4)^54^. When duplicate samples or a pair of samples with cryptic relatedness (PI_HAT >= 0.5) were identified, we removed one sample from the pair. A total of 835 CSF, 529 plasma and 380 brain samples from Washington University passed filters on both genomics and proteomics QC.

### pQTL identification

To test for the association between genetic variants and protein levels we performed a linear regression (additive model), including age, sex, principal component factors from population stratification and genotype platform as covariate:

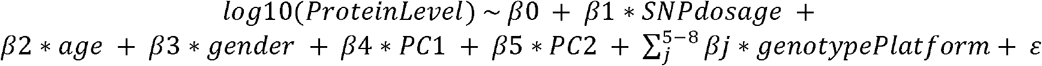

#### cis-pQTL mapping

We conducted cis-pQTL mapping within each of the three tissues. Only proteins passing QC were included in the analyses. Protein level was 10-based logarithm transformed to approximate the normal distribution. Within each tissue, cis-pQTL were identified by linear regression, as implemented in PLINK2 (v2.00a2LM)^54^, adjusting for sex, age, and the first two genotype-based principal components (PCs) and genotyping platforms (Omni1, Omini2.5, NeuroX). We restricted our search to variants within 1 Mb upstream and downstream of the gene body by which each protein was coded. Nominal P-values for each variant–protein pair were estimated using a t-test (two-sided). To identify the list of all significant variant–protein pairs associated with pGenes, variants with a nominal P-value below the genome-wide significance (5×10^−8^) level were considered significant and included in the final list of variants–protein pairs.

#### trans-pQTL mapping

PLINK2 (v2.00a2LM)^54^ was used to test all autosomal variants (MAF > 0.02) using the same QC pipelines as cis-pQTL mapping, but was restricted to variants and proteins encoded by the genes locating outside the 2Mb window, in each tissue independently using an additive linear model. For trans-pQTL mapping, we tested variants for association with the same protein list as for cis-pQTL mapping. We included the covariates of the first two genotype PCs, age, sex, and genotyping platforms when performing association tests. The correlation between variant and protein levels was evaluated using the estimated t-statistic from this model, and list of all significant variant–protein pairs associated with pGenes, variants with a nominal P-value below the genome-wide significance (5×10^−8^/number_PCs) were considered significant and included in the final list of variant–protein pairs. The number of PCs was derived as the minimum principal component number that cumulatively explain 95% of the variance for each tissue after QC. For CSF, plasma and brain, the number of PCs is 169, 230, and 75 respectively. Thus, the p-value thresholds are 3×10^−10^, 2×10^−10^, and 7×10^−10^ respectively.

#### Disease specific analyses

We performed similar analyses including disease status in the model:

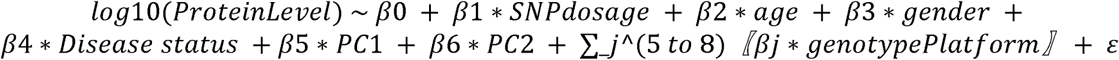

To confirm that none the of findings were driving by disease status.

We also performed analyses sin cases or controls only and compare the β1 for all the analyses to identify any disease-specific effect.

#### Variance explained by the top variant on certain protein

We calculated the adjusted R square value using the linear regression model with the top variant as the only predictor and the log10-based certain protein abundance as the outcome.

### Conditional analysis on locally independent pQTL

To identify conditionally significant associations, we performed a stepwise conditional analysis on all pQTL from round_0 using PLINK2 (v2.00a2LM)^54^ with the –condition or --condition-list option. We used the same significance threshold of P-value = 5×10^−8^ used for the univariable analysis on identifying independent local pQTL.

Conditional analyses were performed as follows: Before conditional (row-1), no SNPs were used as a covariate given one locus. For round-1 (row-2) conditioning, the top SNP from the before-conditioning stage given the same locus was used as an additional covariate in the default model. For round-2 (row-3) conditioning, the top SNP from the before-conditioning stage and the top SNP from round-1 stage were used as an additional covariate in the default model. Both SNPs were within the same locus. This iteration continued for each round by adding one more top SNP from the prior round until no variants passed the genome-wide significance threshold given the same locus. For CSF and plasma samples, in total four rounds of conditional analyses were performed. For brain samples, three rounds of conditional analyses were performed. The result was visualized using locusZoom v1.3^55^.

### Replication strategy for CSF pQTL

To identify all previously reported pQTL from large-scale protein-level GWAS, we performed the CSF replication analyses outline here: We first searched the processed pQTL results using Sasayama et al., 2017 (SOMAscan-based, CSF) and Parkinson’s Progression Markers Initiative (PPMI) released in December 2019, unpublished (SOMAscan-based, CSF) and meta-analysis of these two prior studies; We next checked summary statistics from INTERVAL (SOMAscan-based, plasma). Finally, we queried other plasma pGWAS findings from EBI-NHGRI using phenoscannerV2^56^.

#### Reprocessing pQTL using Sasayama 2017 CSF SOMAscan individual data

To replicate CSF pQTL, we performed linear regression on all proteins using the individual level proteomic and genotype data from Sasayama and colleagues published in 2017 ^11^. We decided to reprocess the pQTL analyses because the original studies used unimputed genotype data. We performed imputation in the Sasyama data in order to have a similar number of SNPs across studies. For proteomics QC, only IQR strategy was used as neither calibrator nor negative control values were provided. Protein outlier was identified if this subject was located outside of either end of distribution using 1.5-fold of IQR given the log10 transformation of the protein level.

Collectively, if the number of outliers given an analyte was greater than 85% of total sample size, this analyte was filtered. Next, an orthogonal approach was used to call subject outliers based on IQR. Collectively, if the number of outliers given an analyte was greater than 85% of total number of analytes passed QC, this subject was labeled as an outlier. Overall 1128 proteins and 133 subjects passed protein data QC. Genotype data QC and imputation was performed as described above. A total of over 5 million (5,187,563) imputed and directly-genotyped SNPs and 154 individuals were used for final analyses. Population substructure analyses was performed as described above. A total of 132 CSF samples from study by Sasayama and colleagues passed filters on both genomics and proteomics QC. We performed linear regression (additive model), including first two principal component factors from population stratification as covariates.

#### Reprocessing pQTL using PPMI 2019 CSF SOMAscan data

To replicate CSF pQTL, we performed linear regression on all shared 709 proteins using the proteomic and genotype data from Parkinson’s Progression Markers Initiative (PPMI) released in December 2019, unpublished. We performed protein QC, genotype imputation and QC and analyses using the same protocols as the ones used for the data generated in this study and described above. A total of over 7 million (7,392,620) imputed and directly-genotyped SNPs and 132 CSF samples from study by PPMI released December 2019 passed filters on both genomics and proteomics QC. We performed a linear regression (additive model), including age, gender and first two principal component factors from population stratification as covariates.

#### Meta-analyses on pQTL using summary statistics from single studies

To replicate more pQTL, we performed fixed effect meta-analyses using METAL^57^. Overall, we performed four different combinations of meta-analyses: 1) meta1_PPMI19_JP17: meta-analysis on both the CSF studies by Sasayama and colleagues published in 2017 and by PPMI released in 2019; 2) meta2_WUcsf_PPMI19_JP17: meta-analysis on all three CSF studies by Sasayama and colleagues published in 2017, by PPMI released in 2019, and by Washington University cohort (this study); 3) meta3_WUcsf_WUplasma_WUbrain: meta-analysis on all three-tissue findings from CSF, plasma and brain respectively by Washington University cohort (this study); 4) meta4_ WUcsf_WUplasma_WUbrain_ PPMI19_JP17: meta-analysis on both the CSF studies by Sasayama and colleagues published in 2017 and by PPMI released in 2019 plus all three-tissue findings from CSF, plasma and brain respectively by Washington University cohort (this study).

### Replication strategy for plasma pQTL

To identify all previously reported pQTL from large-scale protein-level GWAS, we performed CSF replication analyses using the following strategies. We first searched the processed pQTL results using AddNeuroMed (SOMAscan-based, plasma). We next checked summary statistics from INTERVAL (SOMAscan-based, plasma). Finally, we queried other plasma pGWAS findings from EBI-NHGRI using phenoscannerV2.

#### Reprocessing pQTL using AddNeuroMed plasma SOMAscan data

To replicate plasma pQTL, we performed linear regression on all proteins using the proteomic and genotype data from the AddNeuroMed consortium ^23^. A total of over 7 million (7,313,640) imputed and directly-genotyped SNPs and 343 plasma samples from study by AddNeuroMed passed filters on both genomics and proteomics QC. We performed a linear regression (additive model), including age, gender, first two principal component factors from population stratification, centers, status, visit cohorts and batch effects as covariates.

### Replication strategy for brain pQTL

To identify all previously reported pQTL from large-scale protein-level GWAS, we performed the CSF replication analyses using the strategies as follows: we first searched the processed pQTL results using results from our CSF findings; We next queried all plasma pGWAS findings from EBI-NHGRI using phenoscannerV2.

For each locus, we investigated whether the sentinel SNP or a proxy (r^2^ > 0.5) was associated with the same Target protein (or aptamer) in our study at different defined significance thresholds. For the known category in our primary assessment, we used a P-value threshold of 5×10^−8^. For the replicated category in our primary assessment, we used a P-value threshold of 5×10^−2^.

### Overlapping of cis-pQTL and GTEx defined cis-eQTL

For this analysis, we restricted eQTL result reported by GTEx latest and final release v8 ^5^, since this project provided the most comprehensive summary statistics across a wide range of 50 tissue types, including nine brain regions and whole blood. Specifically, the GTEx eQTL results were filtered to contain only cis-variants (also within 1 Mb upstream and downstream) of genes that encode proteins found in our pQTL study and only the common variants in both data sets were used. The metasoft output from multi-tissue analyses^58^ was used for the gene-variant pair extraction, and the beta_fixed effect was used to correlate with pQTL effect size.

### Identification of tissue-specific/shared pQTL

In order to investigate tissue-specificity of pQTL, we performed two steps of analysis. 411 proteins were available all from Washington University dataset. We first assembled a table with P-values for every top pQTL per tissue across all three tissues. We identified the tissue-specificity under three different significant thresholds: 1) a P-value threshold of 5×10^−8^ for all three tissues; 2) a P-value threshold of 5×10^−8^ for one tissue, and 3) a P-value threshold of 5×10^−2^ for the other two tissues. The rationale to perform the 2^nd^ and 3^rd^ analyses with more permissive p-values was because our sample size was not consistent across tissues. As CSF had a higher number of samples and therefore these results could be biased by statistical power, we performed similar analyses with more permissive p-values. We used Venn-diagrams to visualize the tissue specific and shared pQTL. The Venn Diagrams were drawn using VennDiagram^48^ R package.

Second, we curated another table with P-values for every top CSF and brain pQTL from Washington University datasets, plus plasma pQTL from the INTERVAL study that includes more than 3301 samples where 616 proteins were available and performed similar comparisons as we did in step 1. This step was to demonstrate that findings from step-1 were not a product of different sample size.

### Annotation of pQTL

All significant pQTL (hg19) were annotated using ANNOVAR^59^ version (2018-04-16) with the option-geneanno in gene-based annotation mode. Genomic features and variants affecting the nearest genes were used for downstream analyses. The bar plots were drawn using the ggplot2^60^ R package.

### Identification of pleiotropic regions

To identify unique non-overlapping regions associated with a given an aptamer, we first defined a 1-Mb region upstream and downstream of each significant variant for that aptamer. Within the region (2Mb) containing the variant with the smallest P value, any overlapping regions were then merged into the same locus. To identify whether a region was associated with multiple aptamers, we next used an LD-based clumping approach (LD block from the 1000 Genome Project implemented into the ROGHE^61^ R package). Variants were combined into a single region per LD (EUR) defined loci. Any loci associated with more than one protein were identified as pleiotropic regions.

### Investigation of disease status effect on pQTL

To investigate of disease status effect on pQTL, we performed linear regression on the same protein-loci pairs (before conditioning on top variants) identified from the above default model using three additional models: 1) joint analysis including disease status as another covariate (CO vs non-CO); 2) AD case (CA) only using the same covariates as default model; 3) cognitive unimpaired (CO) only using the same covariates as default model. Using scatterplots, we visualized the correlation between each of the additional models and our default model. Using model 1 for comparison, we observed a Pearson correlation coefficient of 0.999 (p-value < 2.2 × 10^−16^), 0.999 (p-value = 4.3 × 10^−202^), 0.999 (p-value = 9.5 × 10^−52^) for CSF, plasma, and brain respectively. Using model 2 for comparison, we observed a Pearson correlation coefficient of 0.991 (p-value = 3.9 × 10^−160^), 0.989 (p-value = 1.8 × 10^−83^), 0.998 (p-value = 2.4 × 10^−29^) for CSF, plasma, and brain respectively. Using model 3 for comparison, we observed a Pearson correlation coefficient of 0.999 (p-value = 5.2 × 10^−234^), 0.998 (p-value = 1.17 × 10^−122^), 0.602 (p-value = 0.002) for CSF, plasma, and brain respectively. The relatively low correlation seen when using model 3 for comparison with controls only in brain samples was due to a much smaller sample size as a control for brain samples.

### Performing MR using TwoSampleMR R package

We used the R package TwoSampleMR v0.4.22^62^, which includes two primary methods. For single SNP, the most basic method, the Wald ratio, was used. For multiple SNPs without pleiotropy, the inverse variance weighted (IVW) estimator was used. This is a meta-analysis of each Wald ratio for each SNP. The regression is constrained to pass through the origin, thus leading to a zero-intercept. This package also implements the harmonization steps before performing MR, and these steps are: 1) Correct the wrong effect/non-effect alleles; 2) Correct the strand issues; 3) Fix the palindromic SNPs; 4) Remove the SNPs with incompatible alleles. The SNPs selected for the analysis were the based on suggestive threshold, 1×10^−5^. The beta-coefficients and standard errors (SEs) for the selected variants (pQTL) from this study were used as input instrumental variables. These instrumental variables were also extracted from the summary statistics from the latest genome-wide association studies for the outcome on neurological disease related traits. (Details see TableS16; Briefly, AD-risk GWAS was published in 2019^3^; AD-progression GWAS in 2018^63^; AD-age at onset GWAS in 2017 ^64^; PD-risk GWAS in 2019^36^; ALS-risk GWAS in 2016^65^; FTD-risk GWAS in 2014^66^; Stroke-risk GWAS in 2018^67^)

### Overlap of proteins with pQTL and drug targets

To obtain information on drugs that target proteins with pQTL from this study, we used the DrugBank database (as of 1/3/2020)^45^. This is a manually curated database that maintains profiles for >15,000 drugs. For our analysis, we focused on the protein target for each drug. For each protein assayed, we identified all drugs in the DrugBank with a matching protein target based on UniProt ID, annotated via https://www.uniprot.org/database/DB-0019. We further integrate the MR analyses result into the overlap of proteins with pQTL and drug targets.

## Data Availability

All data is available at https://ontime.wustl.edu/

## Data availability

All data is available in the NIA-approved National Institute on Aging Genetics of Alzheimer’s Disease Data Storage Site (NIAGADS), the Knight-ADRC (https://knightadrc.wustl.edu/research/resourcerequest.htm), the Center for Neurogenomics and informatics (NGI) website (https://neurogenomics.wustl.edu/). The summary statistics for all the analyses can be easily explored in the ONTIME (Online Neurodegenerative Trait Integrative Multi-Omics Explorer) pheweb (https://ontime.wustl.edu/).

## Acknowledgments

We thank all the participants and their families, as well as the many institutions and their staff.

## Funding

This work was supported by grants from the National Institutes of Health (R01AG044546, P01AG003991, RF1AG053303, R01AG058501, U01AG058922, RF1AG058501 and R01AG057777), the Alzheimer Association (NIRG-11-200110, BAND-14-338165, AARG-16-441560 and BFG-15-362540). This work was supported by access to equipment made possible by the Hope Center for Neurological Disorders, and the Departments of Neurology and Psychiatry at Washington University School of Medicine. The recruitment and clinical characterization of research participants at Washington University were supported by NIH P50 AG05681, P01 AG03991, and P01 AG026276.

## Author contributions

CY performed the analyses, interpreted the results, and wrote the manuscript. FGF, LI, MVF, FW, JLB, ZL, UD, KM and JPB contributed to data collection, data processing, QC, and cleaning. JCM, AF, RJP contributed samples and/or data. BS wrote the manuscript. JAB and OH developed the pheweb browser. HR, BB, OH and CC designed the study, collected the data, supervised the analyses, interpreted the results, and wrote the manuscript. All authors read and contributed to the final manuscript.

## Competing interests

CC receives research support from: Biogen, EISAI, Alector and Parabon. The funders of the study had no role in the collection, analysis, or interpretation of data; in the writing of the report; or in the decision to submit the paper for publication. CC is a member of the advisory board of Vivid genetics, Halia Therapeutics and ADx Healthcare.

## Materials & Correspondence

Materials and correspondence should be addressed Carlos Cruchaga.

## Notes

### Author Declarations

The Institutional Review Boards of Washington University School of Medicine in St. Louis approved the study, and research was carried out in accordance with the approved protocols.

## References

1. Altshuler, D., Daly, M. J. & Lander, E. S. Genetic Mapping in Human Disease. Science 322, 881–888 (2008).

2. Morris, A. P. et al. Large-scale association analysis provides insights into the genetic architecture and pathophysiology of type 2 diabetes. Nature Genetics 44, 981–990 (2012).

3. Kunkle, B. W. et al. Genetic meta-analysis of diagnosed Alzheimer’s disease identifies new risk loci and implicates Aβ, tau, immunity and lipid processing. Nature Genetics 51, 414 (2019).

4. Claussnitzer, M. et al. A brief history of human disease genetics. Nature 577, 179–189 (2020).

5. Aguet, F. et al. The GTEx Consortium atlas of genetic regulatory effects across human tissues. bioRxiv 787903 (2019) doi:10.1101/787903.

6. van der Wijst, M. G. P. et al. Single-cell eQTLGen Consortium: a personalized understanding of disease. 1909.12550 [q-bio] (2019).

7. Sun, B. B. et al. Genomic atlas of the human plasma proteome. Nature 558, 73–79 (2018).

8. Suhre, K. et al. Connecting genetic risk to disease end points through the human blood plasma proteome. Nature Communications 8, 14357 (2017).

9. Folkersen, L. et al. Mapping of 79 loci for 83 plasma protein biomarkers in cardiovascular disease. PLOS Genetics 13, e1006706 (2017).

10. Deming, Y. et al. Genetic studies of plasma analytes identify novel potential biomarkers for several complex traits. Scientific Reports 6, 18092 (2016).

11. Sasayama, D. et al. Genome-wide quantitative trait loci mapping of the human cerebrospinal fluid proteome. Hum Mol Genet 26, 44–51 (2017).

12. Kauwe, J. S. K. et al. Genome-Wide Association Study of CSF Levels of 59 Alzheimer’s Disease Candidate Proteins: Significant Associations with Proteins Involved in Amyloid Processing and Inflammation. PLOS Genetics 10, e1004758 (2014).

13. Robins, C. et al. Genetic control of the human brain proteome. bioRxiv 816652 (2019) doi:10.1101/816652.

14. Aguet, F. et al. Genetic effects on gene expression across human tissues. Nature 550, 204–213 (2017).

15. Hillary, R. F. et al. Genome and epigenome wide studies of neurological protein biomarkers in the Lothian Birth Cohort 1936. Nat Commun 10, 3160–3160 (2019).

16. Yao, C. et al. Genome[wide mapping of plasma protein QTLs identifies putatively causal genes and pathways for cardiovascular disease. Nature Communications 9, 3268 (2018).

17. Paré, G. et al. Novel association of ABO histo-blood group antigen with soluble ICAM-1: results of a genome-wide association study of 6,578 women. PLoS Genet. 4, e1000118 (2008).

18. Lovestone, S. et al. AddNeuroMed—The European Collaboration for the Discovery of Novel Biomarkers for Alzheimer’s Disease. Annals of the New York Academy of Sciences 1180, 36–46 (2009).

19. Boyle, A. P. et al. Annotation of functional variation in personal genomes using RegulomeDB. Genome Res. 22, 1790–1797 (2012).

20. Jayaratnam, S., Khoo, A. K. L. & Basic, D. Rapidly progressive Alzheimer’s disease and elevated 14-3-3 proteins in cerebrospinal fluid. Age Ageing 37, 467–469 (2008).

21. Foote, M. & Zhou, Y. 14-3-3 proteins in neurological disorders. Int J Biochem Mol Biol 3, 152–164 (2012).

22. Ibanez, L. et al. Overlap in the Genetic Architecture of Stroke Risk, Early Neurological Changes, and Cardiovascular Risk Factors. Stroke 50, 1339–1345 (2019).

23. Lourdusamy, A. et al. Identification of cis-regulatory variation influencing protein abundance levels in human plasma. Hum Mol Genet 21, 3719–3726 (2012).

24. Walker, R. L. et al. Genetic Control of Expression and Splicing in Developing Human Brain Informs Disease Mechanisms. Cell 179, 750-771.e22 (2019).

25. Orozco, L. D. et al. Integration of eQTL and a Single-Cell Atlas in the Human Eye Identifies Causal Genes for Age-Related Macular Degeneration. Cell Reports 30, 1246-1259.e6 (2020).

26. Ndungu, A., Payne, A., Torres, J. M., Bunt, M. van de & McCarthy, M. I. A Multi-tissue Transcriptome Analysis of Human Metabolites Guides Interpretability of Associations Based on Multi-SNP Models for Gene Expression. The American Journal of Human Genetics 0, (2020).

27. Del-Aguila, J. L. et al. A single-nuclei RNA sequencing study of Mendelian and sporadic AD in the human brain. Alzheimer’s Research & Therapy 11, 71 (2019).

28. Jonsson, T. et al. Variant of TREM2 associated with the risk of Alzheimer’s disease. N. Engl. J. Med. 368, 107–116 (2013).

29. Guerreiro, R. et al. TREM2 variants in Alzheimer’s disease. N. Engl. J. Med. 368, 117–127 (2013).

30. Griciuc, A. et al. TREM2 Acts Downstream of CD33 in Modulating Microglial Pathology in Alzheimer’s Disease. Neuron 0, (2019).

31. Chan, G. et al. CD33 modulates TREM2: convergence of Alzheimer loci. Nature Neuroscience 18, 1556–1558 (2015).

32. Deming, Y. et al. The MS4A gene cluster is a key modulator of soluble TREM2 and Alzheimer’s disease risk. Science Translational Medicine 11, eaau2291 (2019).

33. Cruchaga, C. et al. Cerebrospinal fluid APOE levels: an endophenotype for genetic studies for Alzheimer’s disease. Hum Mol Genet 21, 4558–4571 (2012).

34. Nalls, M. A. et al. Large-scale meta-analysis of genome-wide association data identifies six new risk loci for Parkinson’s disease. Nat. Genet. 46, 989–993 (2014).

35. Chang, D. et al. A meta-analysis of genome-wide association studies identifies 17 new Parkinson’s disease risk loci. Nat. Genet. 49, 1511–1516 (2017).

36. Nalls, M. A. et al. Identification of novel risk loci, causal insights, and heritable risk for Parkinson’s disease: a meta-analysis of genome-wide association studies. The Lancet Neurology 18, 1091–1102 (2019).

37. Krohn, L. et al. Genetic, Structural, and Functional Evidence Link TMEM175 to Synucleinopathies. Ann. Neurol. 87, 139–153 (2020).

38. Witoelar, A. et al. Genome-wide Pleiotropy Between Parkinson Disease and Autoimmune Diseases. JAMA Neurol 74, 780–792 (2017).

39. Hook, P. W. et al. Single-Cell RNA-Seq of Mouse Dopaminergic Neurons Informs Candidate Gene Selection for Sporadic Parkinson Disease. Am. J. Hum. Genet. 102, 427–446 (2018).

40. Poletto, E. et al. Effects of gene therapy on cardiovascular symptoms of lysosomal storage diseases. Genetics and Molecular Biology 42, 261–285 (2019).

41. Alector Inc. First in Human Study for Safety and Tolerability of AL003. - Full Text View - ClinicalTrials.gov. https://clinicaltrials.gov/ct2/show/NCT03822208.

42. Liu, X., Lu, D., Bowser, R. & Liu, J. Expression of Carbonic Anhydrase I in Motor Neurons and Alterations in ALS. International Journal of Molecular Sciences 17, 1820 (2016).

43. Lu, D. et al. Carbonic Anhydrase I modifies SOD1-induced motor neuron toxicity in <em>Drosophila</em> via ER stress pathway. Journal of Neuroscience and Neurological Disorders 3, 135–144 (2019).

44. Bethea, J. W. Clinical Anesthesia, 6th Edition. Anesthesiology 112, 767–768 (2010).

45. Wishart, D. S. et al. DrugBank: a comprehensive resource for in silico drug discovery and exploration. Nucleic Acids Res 34, D668–D672 (2006).

46. Cohlberg, J. A., Li, J., Uversky, V. N. & Fink, A. L. Heparin and other glycosaminoglycans stimulate the formation of amyloid fibrils from alpha-synuclein in vitro. Biochemistry 41, 1502–1511 (2002).

47. Holmes, B. B. et al. Heparan sulfate proteoglycans mediate internalization and propagation of specific proteopathic seeds. Proc. Natl. Acad. Sci. U.S.A. 110, E3138–3147 (2013).

48. Chen, H. VennDiagram: Generate High-Resolution Venn and Euler Plots. (2018).

49. Morris, J. C. The Clinical Dementia Rating (CDR): current version and scoring rules. Neurology 43, 2412–2414 (1993).

50. Gold, L. et al. Aptamer-Based Multiplexed Proteomic Technology for Biomarker Discovery. PLOS ONE 5, e15004 (2010).

51. Sattlecker, M. et al. Alzheimer’s disease biomarker discovery using SOMAscan multiplexed protein technology. Alzheimers Dement 10, 724–734 (2014).

52. Williams, S. A. et al. Plasma protein patterns as comprehensive indicators of health. Nature Medicine 1–7 (2019) doi:10.1038/s41591-019-0665-2.

53. UniProt: a worldwide hub of protein knowledge. Nucleic Acids Res 47, D506–D515 (2019).

54. Chang, C. C. et al. Second-generation PLINK: rising to the challenge of larger and richer datasets. GigaScience 4, 7 (2015).

55. Pruim, R. J. et al. LocusZoom: regional visualization of genome-wide association scan results. Bioinformatics 26, 2336–2337 (2010).

56. Kamat, M. A. et al. PhenoScanner V2: an expanded tool for searching human genotype–phenotype associations. Bioinformatics (2019) doi:10.1093/bioinformatics/btz469.

57. Willer, C. J., Li, Y. & Abecasis, G. R. METAL: fast and efficient meta-analysis of genomewide association scans. Bioinformatics 26, 2190–2191 (2010).

58. Sul, J. H., Han, B., Ye, C., Choi, T. & Eskin, E. Effectively identifying eQTLs from multiple tissues by combining mixed model and meta-analytic approaches. PLoS Genet. 9, e1003491 (2013).

59. Wang, K., Li, M. & Hakonarson, H. ANNOVAR: functional annotation of genetic variants from high-throughput sequencing data. Nucleic Acids Res 38, e164–e164 (2010).

60. Wickham, H. ggplot2: Elegant Graphics for Data Analysis. (Springer-Verlag, 2009). doi:10.1007/978-0-387-98141-3.

61. Mancuso, N. et al. Integrating Gene Expression with Summary Association Statistics to Identify Genes Associated with 30 Complex Traits. The American Journal of Human Genetics 100, 473–487 (2017).

62. Hemani, G. et al. The MR-Base platform supports systematic causal inference across the human phenome. eLife 7, e34408 (2018).

63. Del-Aguila, J. L. et al. Assessment of the Genetic Architecture of Alzheimer’s Disease Risk in Rate of Memory Decline. J. Alzheimers Dis. 62, 745–756 (2018).

64. Huang, K. et al. A common haplotype lowers PU.1 expression in myeloid cells and delays onset of Alzheimer’s disease. Nature Neuroscience 20, 1052–1061 (2017).

65. van Rheenen, W. et al. Genome-wide association analyses identify new risk variants and the genetic architecture of amyotrophic lateral sclerosis. Nature Genetics 48, 1043–1048 (2016).

66. Ferrari, R. et al. Frontotemporal dementia and its subtypes: a genome-wide association study. Lancet Neurol 13, 686–699 (2014).

67. Malik, R. et al. Multiancestry genome-wide association study of 520,000 subjects identifies 32 loci associated with stroke and stroke subtypes. Nature Genetics 50, 524 (2018).

